# Evaluating High-Frequency Automated Cognitive Tasks Across Immune-Mediated Inflammatory Disease and Neurodegenerative Disease Patients

**DOI:** 10.64898/2026.04.26.26351685

**Authors:** Alexander J Kaula, Nick Taptiklis, Francesca Cormack, Laura C. M. Kuijper, Stefan Avey, Meenakshi Chatterjee, Rana Zia Ur Rehman, Susanne de Bot, Andrea Pilotto, C Janneke van der Woude, Christopher A Lamb, Ralf Reilmann, Nikolay V. Manyakov, Walter Maetzler, Wan-Fai Ng

## Abstract

This analysis evaluates the feasibility and psychometric properties of daily digital cognitive assessments (DCAs) delivered on smartphones using data from the large, international Identifying Digital Endpoints to Assess FAtigue, Sleep and acTivities of daily living in Neurodegenerative disorders and Immune-mediated inflammatory diseases (IDEA-FAST) study. The data we analyse were collected from patients with neurodegenerative diseases (NDDs) and immune-mediated inflammatory diseases (IMIDs), and healthy controls (a subset who participated in all phases of the study, total N=1005) in their own homes. These data were obtained alongside data from other devices that monitored physiology, kinematics, and sleep quality. Following a baseline visit, participants were remotely monitored via three scheduled daily sessions for 6-7 days in each of 4 active assessment phases (APs). APs were separated by 6-week intervals. Daily schedules comprised a morning psychomotor vigilance task (PVT) with eDiary, afternoon session (eDiary only), and an evening digit symbol substitution task (DSST) with eDiary. We evaluated session coverage using logistic mixed effects, test-retest reliability using ICCs, disease impacts on performance using linear mixed effect ANCOVA, and familiarisation using linear mixed effects.

Overall coverage was 66.7% for the PVT and 76.2% for the DSST, with no significant differences between the healthy volunteers and disease cohorts. Coverage varied significantly by time-of-day (Evening > Morning > Afternoon), and improved with age, with an interaction revealing session time-of-day affected older participants less, all *p* < .001. Coverage was highest in AP 1 and reduced in subsequent APs. AP-day effects on coverage interacted significantly with AP, with a modest decline over AP 1, and the pattern reversed in APs 2-4.

Baseline reliability was good (> .70) for both PVT mean reaction time and DSST total correct across all cohorts, and the movement-based measure from the DSST ranged [.54, .82], with lower values in the Parkinson’s Disease and Primary Sjögren’s Syndrome cohorts.

Both tasks showed significant cohort effects, with performance in IMID cohorts intermediate between healthy controls and NDD. Longitudinal analysis revealed significant familiarisation effects in DSST. This was greatest in healthy controls, with significant attenuation of these effects in disease cohorts. No effect of familiarisation was seen in the PVT.

Collectively, these results support the usefulness of at-home cognitive assessment on smartphones. Brief measures of cognition can be captured remotely in disease as well as controls with good adherence and sensitivity to distinguish known patient groups from healthy controls.

## 1. Introduction

People with neurodegenerative disorders (NDD) and immune-mediated inflammatory diseases (IMID) often show deficits across a range of neuropsychological measures (Whitehouse et al., 2019), alongside worse health-related quality of life (HRQoL), sleep disturbances, and increased fatigue (Enns et al., 2018; Suh, Lee, Kim, & Boo, 2022). The IDEA-FAST Clinical Observational Study^1^is a European multinational study that recruited 2000 participants across NDD and IMID cohorts to identify digital technologies that can support remote monitoring of these conditions (Antikainen et al., 2022).

Cognitive problems are frequently reported as an important component of fatigue. Often informally termed “brain fog”, these problems can encompass subjective difficulties with concentration, memory, thinking and word-finding (Alim-Marvasti et al., 2024). Objective changes in cognitive task performance have also been associated with brain fog, including deficits in attention, short-term and working memory, executive function and speed of processing (Cockshell & Mathias, 2010). However, these findings have not led to a consensus definition or measurement approach, and research has largely examined brain fog within single therapeutic areas such as chronic fatigue syndrome and long COVID rather than across conditions. Although cognitive problems are relatively well studied in NDD, fewer data are available in IMID, and the basis of cognitive impairment in these populations remains less well understood.

Cognitive problems associated with NDD and IMID are often assessed using in-clinic assessments and patient-reported outcomes (PROs) (see, for example, Gwinnut et al., 2022, for a meta-analysis in IMID), but these approaches have important limitations. PROs are prone to recall and response bias and are typically assessed retrospectively over one to two weeks (Stone et al., 2002). Moreover, the relationship between PROs and cognitive function can be confounded by depressive symptoms or poor sleep quality (Siciliano et al., 2021). By assessing cognition in everyday settings, repeated remote digital cognitive assessment offers the promise of overcoming these limitations.

The value of repeated remote assessment depends on the extent to which assessments are successfully completed, and on the psychometric performance of the assessments themselves. Participants must be willing and able to follow the assessment schedule, but frequent testing may increase burden and reduce coverage, particularly in patient groups whose symptoms may interfere with their ability to participate. Coverage may also be affected by participant-level and protocol-level factors, making it important to characterise not only whether coverage differs between healthy and clinical groups, but also which factors shape completion of scheduled assessments. For repeated assessment data to be useful, task performance must be reliable over time and potential familiarisation effects with repeated administration must be understood. Tasks must also be sensitive to differences between health and disease states.

Given this background, the present study examines coverage and psychometric properties of the digital CANTAB cognitive assessments used in the IDEA-FAST study (Maetzler et al., 2026). Specifically, we address three questions. First, what participant- and protocol-level factors influence coverage of remote cognitive assessments, and does coverage differ between healthy individuals and those living with chronic neurological or immune-mediated inflammatory conditions? Second, are smartphone-based measures of psychomotor vigilance and processing speed psychometrically suitable for repeated use in clinical research? Third, do brief, repeatable smartphone cognitive tasks show sensitivity to group-level differences in cognitive performance between healthy and clinical cohorts?

## 2. Methods

### 2.1. Study Design and Participants

The IDEA-FAST multi-centre clinical observational study (COS) enrolled participants over 24 weeks. Participants were recruited from 17 centres across 10 countries within Europe. We analyse data from completed participants who did not declare dropping out. Each participant attended two study visits at the recruitment centre (at week 0 and at the end of the study) as well as two remote visits at week 8 and 16. During study visits, participants completed clinical assessments and various patient-reported outcomes (PROs). Following each visit, participants began an assessment phase (AP) detailed below, the first being 10 days, including three onboarding days, with second, third, and fourth APs being seven days, each including one initial onboarding day. This gives 25 days during which we analyse performance, out of a maximum of 31 (i.e. 10 + 7 + 7 + 7) days’ data.

Participants are enrolled into seven cohorts (see Table 1 for details): healthy volunteers (HV), and six disease cohorts comprising neurodegenerative disorders (NDD) and immune-mediated inflammatory disorders (IMID). NDD cohorts are Huntington’s disease (HD) and Parkinson’s disease (PD). IMID are inflammatory bowel disease (IBD), Primary Sjögren’s Syndrome (PSS, also known as Sjögren’s Disease), rheumatoid arthritis (RA), and systemic lupus erythematosus (SLE).

**Table 1:**
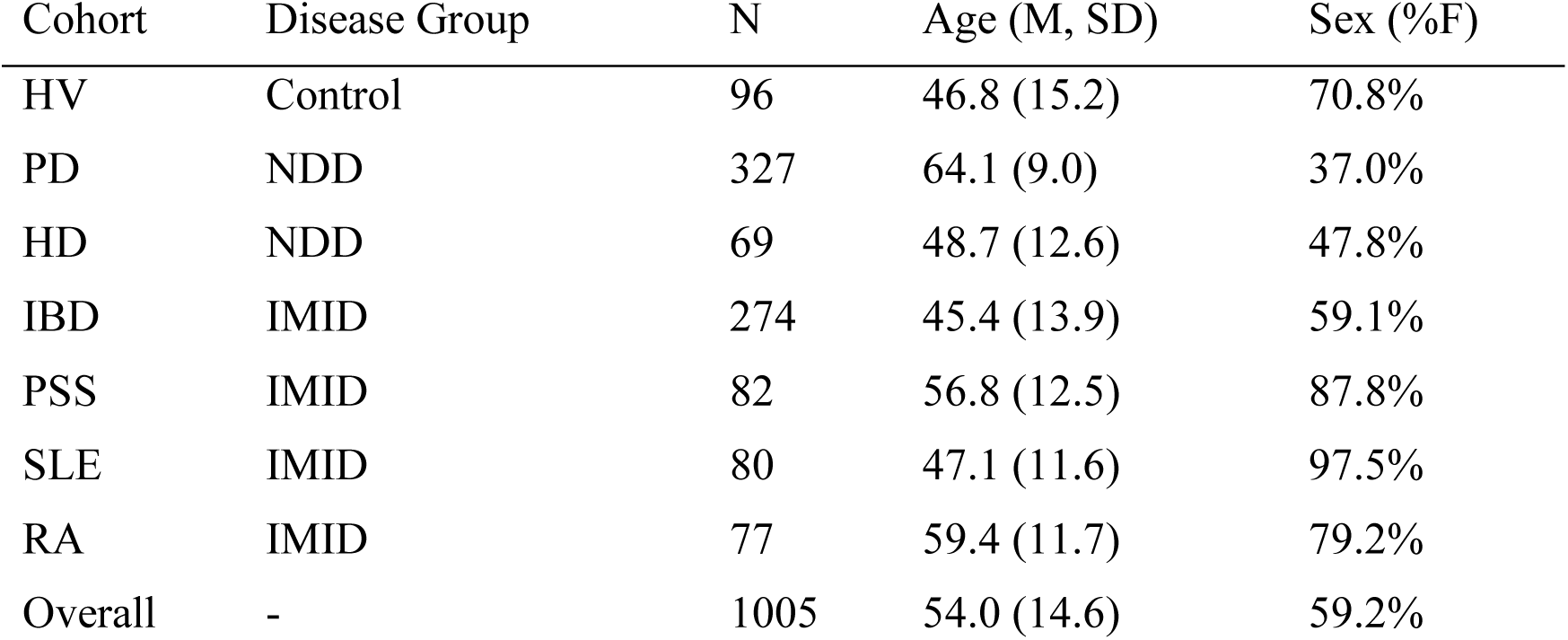
Demographic and Clinical Characteristics of the Study Cohorts.

| Cohort | Disease Group | N | Age (M, SD) | Sex (%F) |
| --- | --- | --- | --- | --- |
| HV | Control | 96 | 46.8 (15.2) | 70.8% |
| PD | NDD | 327 | 64.1 (9.0) | 37.0% |
| HD | NDD | 69 | 48.7 (12.6) | 47.8% |
| IBD | IMID | 274 | 45.4 (13.9) | 59.1% |
| PSS | IMID | 82 | 56.8 (12.5) | 87.8% |
| SLE | IMID | 80 | 47.1 (11.6) | 97.5% |
| RA | IMID | 77 | 59.4 (11.7) | 79.2% |
| Overall | - | 1005 | 54.0 (14.6) | 59.2% |

Inclusion criteria are age of more than 18 years, ability to understand study instructions, and ability to carry out procedures of the study according to the opinion of the investigator (see Maetzler et al., 2026 for full COS protocol).

### 2.2. Assessment Phases

An onboarding telephone call cued participants to begin. To give participants ample opportunity to familiarise themselves with the variety of digital assessments in use, the first three days of the first AP had three designated ‘familiarisation days’, before seven days of data collection. Subsequent APs each began with one familiarisation day. This makes 25 days of total active APs for each participant.

During each AP, participants used smartphones (either theirs, or provisioned) to complete sessions scheduled by the CANTAB App, Cambridge Cognition, 2019). The daily schedule of assessments is shown in Table 2 (below). Participants are initially prompted one hour after the morning, afternoon and evening assessment window opens, with further reminders hourly after that, if no session is completed.

**Table 2:**
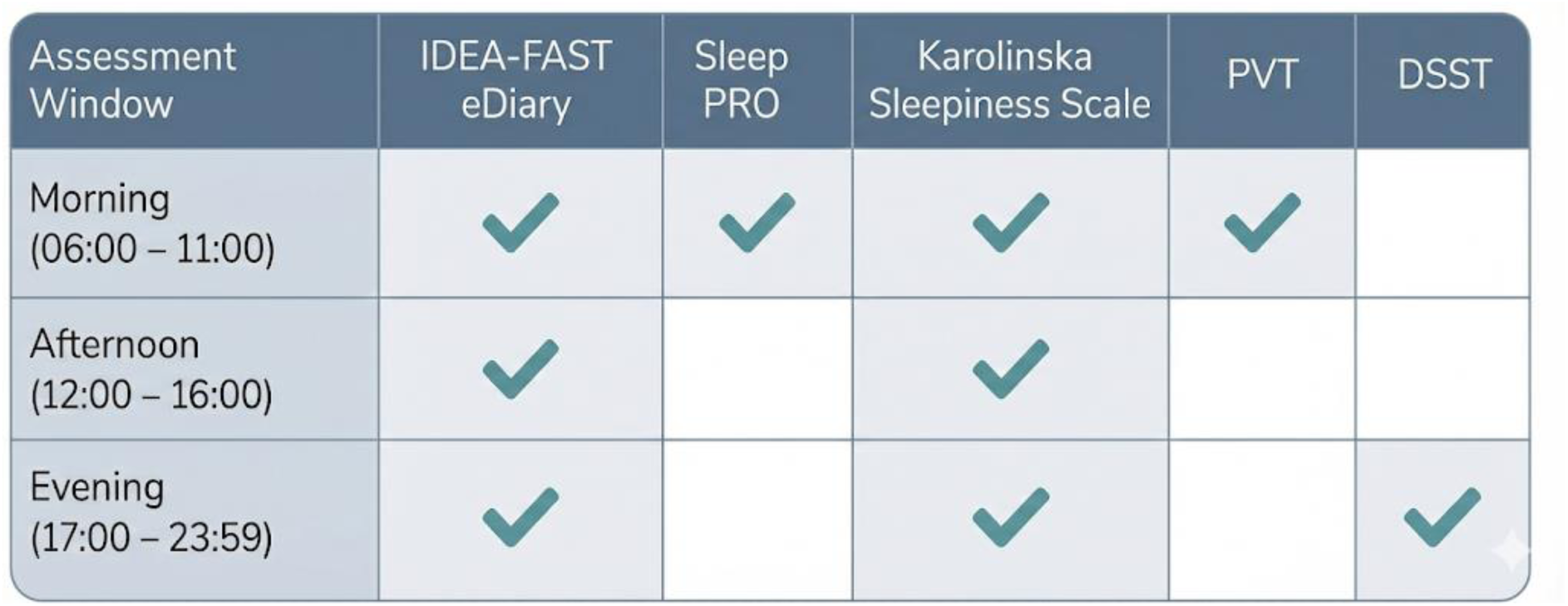
Schedule of assessments within each AP

#### 2.2.1. Daily Patient-Reported Outcomes (PROs)

PRO measures of fatigue (physical, mental, and overall), mood, anxiety, pain (the “IDEA-FAST eDiary”) as well as quality of sleep were recorded using a VAS (scored from 0-100). Present sleepiness was captured using the Karolinska Sleepiness Scale (Åkerstedt & Gilbert, 1990), scored from 0 to 9.

#### 2.2.2. Psychomotor Vigilance Task (PVT)

PVT is considered a test of sustained attention and has been shown particularly sensitive to sleep disruption. Although the standard version of PVT (Dinges & Powell, 1984) is 10 minutes duration, three-minute, shortened versions such as the one in use here (illustrated in left panel, Figure 2, below) demonstrate sensitivity to sleep disruption and are now in common use (for example see Grant et al. 2017). The participant observes an empty rectangle in the entire screen. Following a random delay of between two and six seconds a number appears in the rectangle and begins counting upwards in milliseconds within this rectangle.

The user should tap the screen as quickly as possible after the number appears. If a response is not recorded within 500ms, the response is classed as a lapse. Total responses, lapses, false alarms, and reaction time for correct responses are captured. The key outcome measure is mean reaction time over the task (ms).

#### 2.2.3. Digit Symbol Substitution Test (DSST)

DSST is considered a test of global cognition because it recruits a diverse array of cognitive operations (see McIntyre et al., 2022). In this electronic version of the task (CognitionKit DSST, shown in Panel B Figure 1 below) the participant is shown a digit from one to nine and must match this digit to a corresponding symbol by drawing it using the five-point grid in the space below, aiming to complete as many as possible in a 90-second timed run. A key is displayed at the top of the screen, indicating which digit corresponds to which symbol. This key remains always onscreen. Below the current problem is the subject’s response area. When a digit is presented, the subject must respond by drawing the corresponding symbol using their finger on the five-point grid. The next problem is presented once the subject has responded to the current problem. Responses are scored, {correct, incorrect}, time between the presentation of the target digit and the participant touching the screen (response latency, ms), and the time taken by the participant to draw their correct response once their finger has touched the screen (movement duration, ms) are all recorded. Here, we report the total correct responses (shows as DSTTC on proprietary reports) and the mean correct movement duration (DSTCMLM on reports).

**Figure 1:**
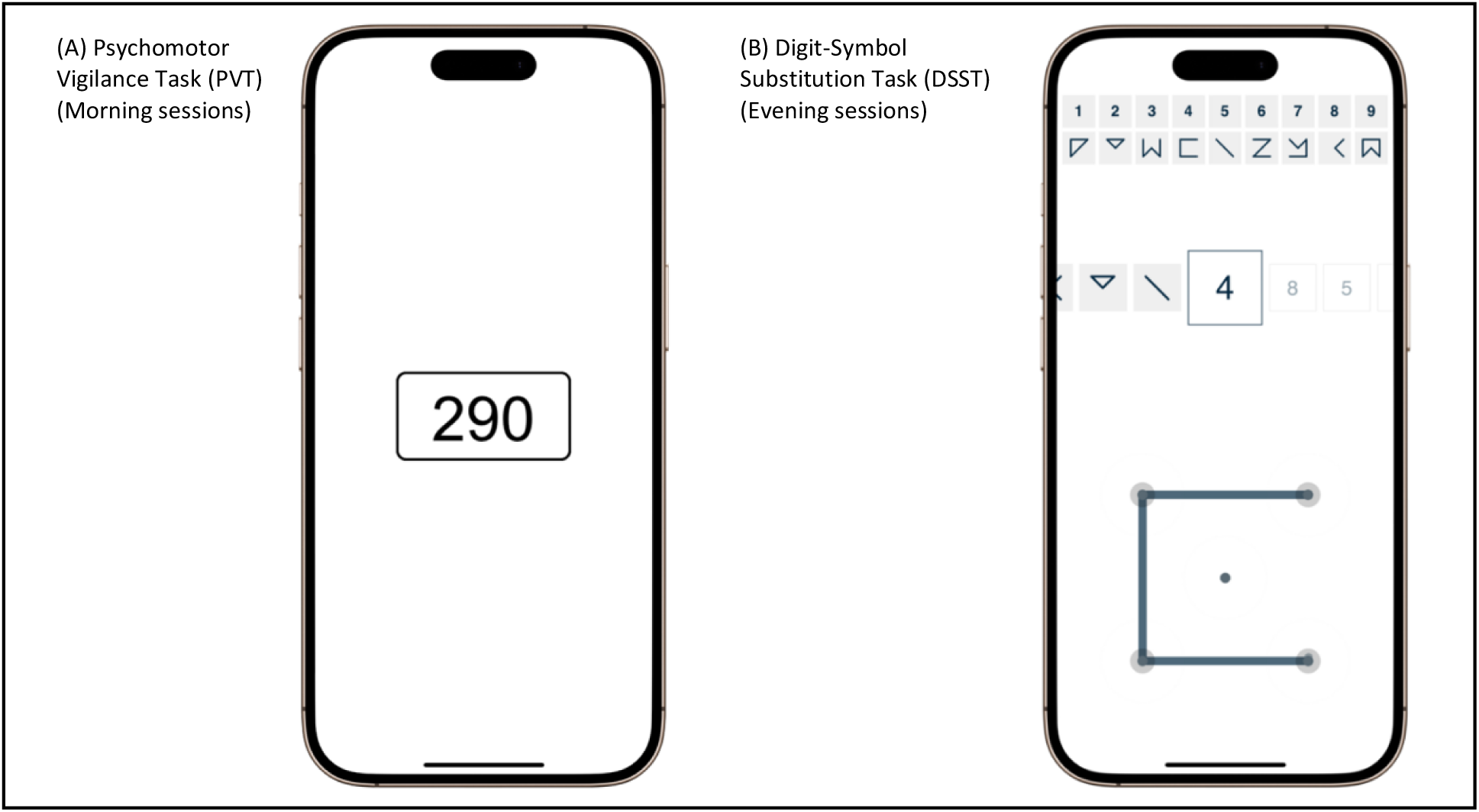
Appearance of the cognitive assessments used in the study, prompted by an app during morning (PVT) and evening (CognitionKit DSST) sessions.

**Figure 2:**
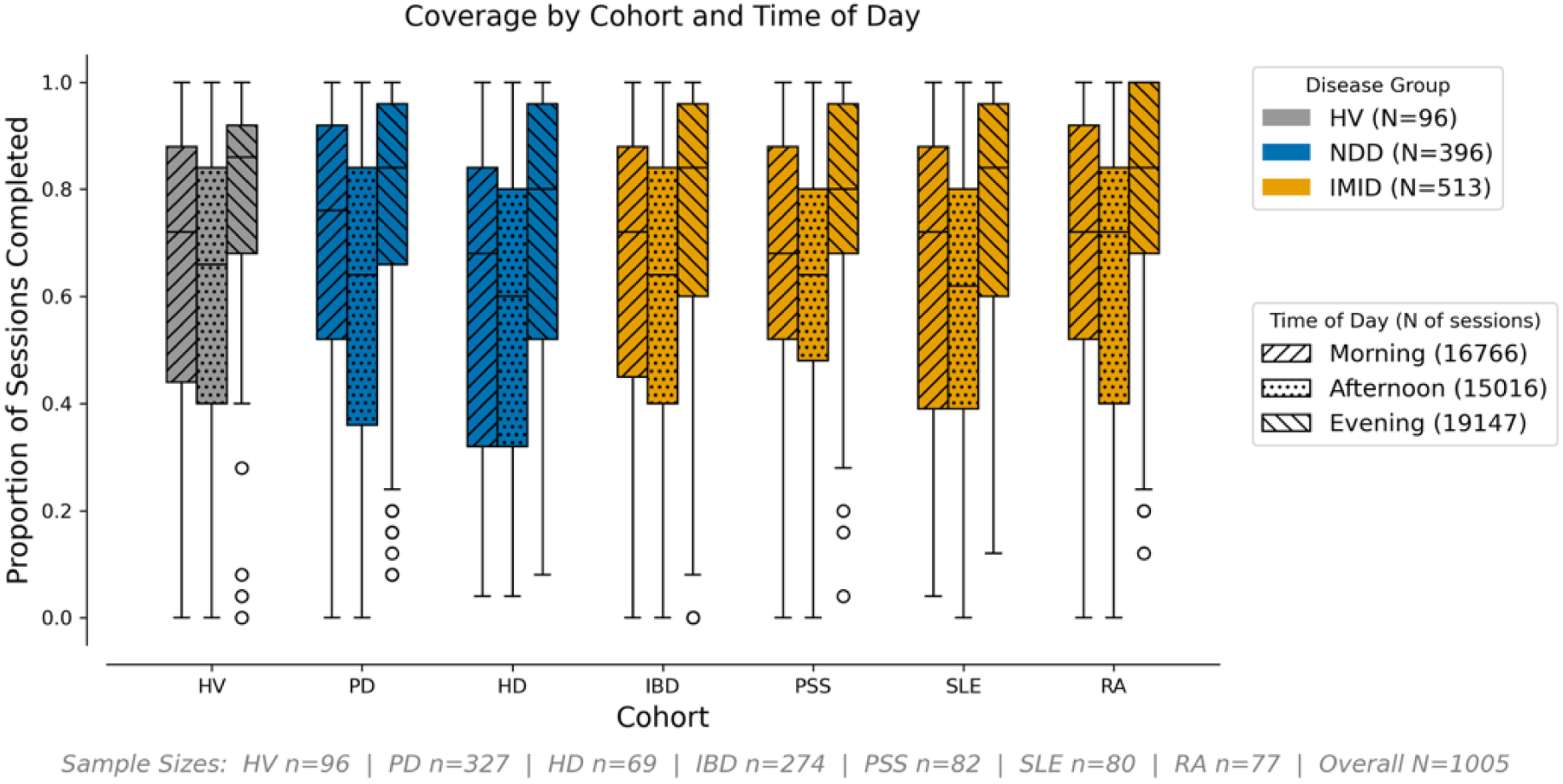
Coverage by cohort and time of day. Boxplots depict the distribution of per-subject completion proportions for morning, afternoon, and evening sessions across the seven cohorts. Shading indicates disease-group membership (grey = HV, blue = neurodegenerative diseases, orange = immune-mediated inflammatory diseases); hatching denotes session time-of-day (diagonal = morning, dotted = afternoon, reverse diagonal = evening). Medians are shown in black. The logistic mixed effects model of coverage showed significant time-of-day (p < .001) effects, with highest adherence in the evening, intermediate in the morning, and lowest in the afternoon. As can be seen in this plot, the schedule / time-of-day effect was consistent over cohorts, and no cohort-level differences from controls survive Holm correction for multiple comparisons.

### 2.3. Analysis

#### 2.3.1. Coverage

##### 2.3.1.1. Definition and Inclusion

Observed coverage was calculated per-subject over the whole study. We excluded AP onboarding days (see above) from our analyses. Thus 25 sessions (one AP of seven days and three APs of six days) were expected per time-period (mornings, afternoons, and evenings) per enrolled participant.

##### 2.3.1.2. Modelling Daily Session Completion Binary Over Study Course

Session completion was analysed as a binary outcome using a logistic mixed-effects model with fixed effects for cohort, z-scored age, session time-of-day (sTOD), AP number (1–4), and AP-day. AP-day was normalized (0 to 1) to account for varying AP lengths. We included age × sTOD and AP × AP-day interactions, with participants and sites as random intercepts. The model intercept represents a Healthy Volunteer at mean age in a morning session midway through AP 1. For interpretability, results are expressed as model-predicted probabilities using the inverse-logit transform, marginalized over other factors via the emmeans package in R (Lenth & Piaskowski, 2025), with differences in odds ratios assessed using Wald’s Z-tests.

#### 2.3.2. Reliability

Reliability of PVT mean reaction time, DSST total correct, and DSST correct movement duration was assessed using two-way mixed effects intraclass correlations for consistency, ICC(3,1). To ensure that estimates reflect measurement precision rather than long-term clinical change, analysis was restricted to the first assessment phase, AP1. Estimates include 95% confidence intervals. Disease cohorts were compared to controls.

#### 2.3.3. Sensitivity to Disease

To assess the sensitivity of the selected measures of PVT and DSST performance to disease, we considered per-subject means over the whole study in a linear mixed-effects ANCOVA with age as a covariate. In post-hoc t-tests we compare age-adjusted means of each cohort against the HV group and use Holm’s correction to control Family Wise Error. We report cohort differences from HV.

#### 2.3.4. Familiarisation Effects on Performance

We assessed the effects of repeated task usage over the whole study by using linear mixed effect (LME) models to model performance as a function of study day (continuous) and cohort (fixed effects), with participant as a random effect to account for repeated measures. To test whether clinical cohorts changed at different rates from controls we included a study-day × Cohort interaction term, adjusting for age. Parameter estimates are reported as a daily rate of change, and the projected change over the course of the 25 study-day period, with Wald’s Z-test comparing modelled cohort slopes, Holm-corrected.

## 3. Results

### 3.1. Coverage

#### 3.1.1. Model-Based Analysis of Session Completion Odds

The observed pattern of adherence to sessions in morning, afternoon, evening, and overall, is described in Table 3. Adherence is highest in AP1, dropping for AP 2 and remaining around the same level inAPs 3, and 4. Adherence is consistently highest for evening sessions, lower for morning sessions, and lowest for afternoon sessions. Figure 2 illustrates the consistent pattern of adherence between cohorts and over session time-of-day. Figure 3 highlights that pattern of adherence over time of day over APs, and the raster in Figure 4 illustrates participation over the course of the study by all participants who remained enrolled over 4 APs.

**Figure 3:**
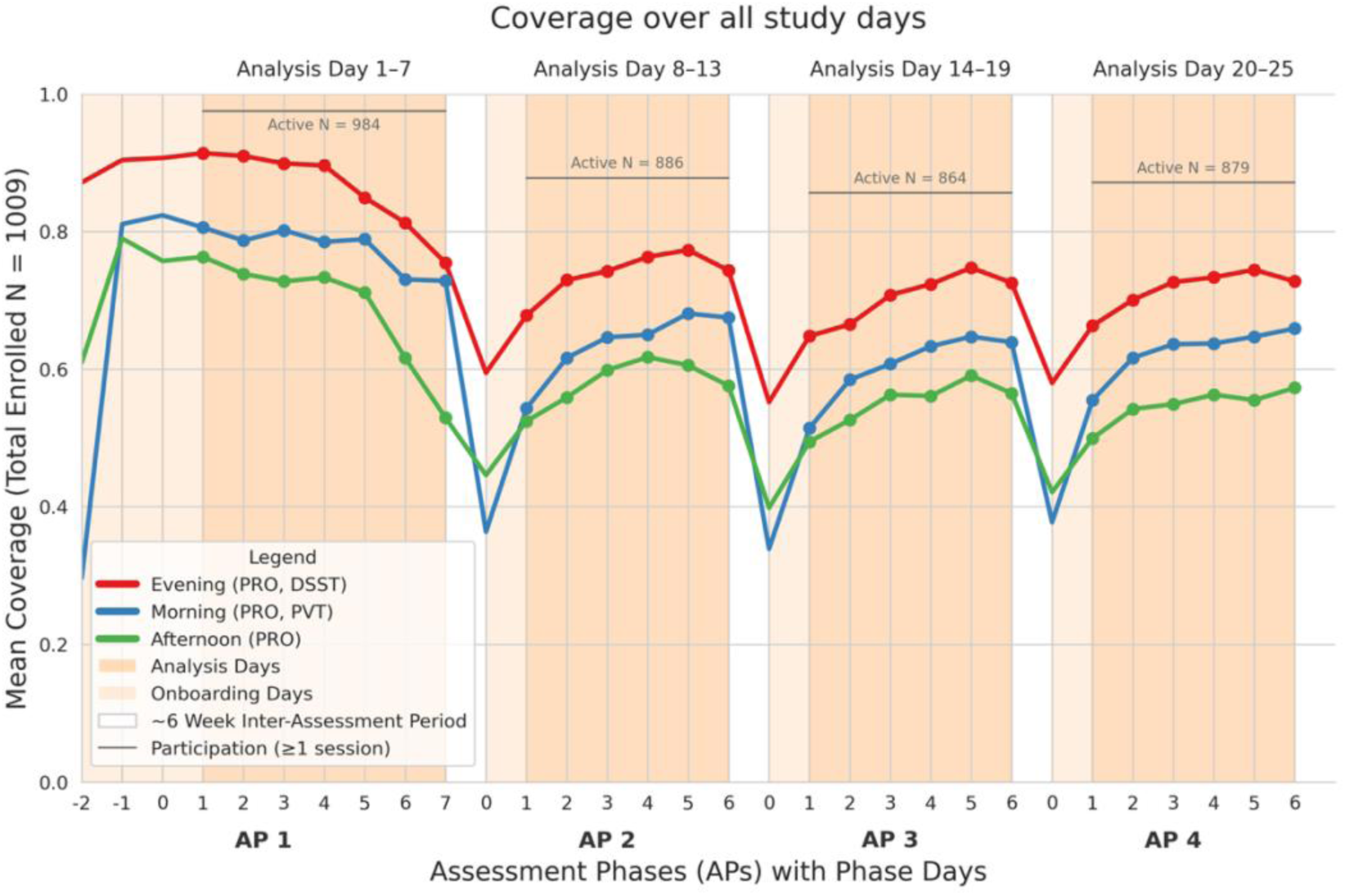
Mean coverage by session period across participants enrolled throughout four APs (approx. 28 weeks). Shaded sections indicate days included in the analysis; paler shading indicates excluded familiarisation periods. Non-shaded sections indicate breaks between APs. During morning sessions, participants completed a PRO questionnaire (QST) and PVT; during the afternoon, the QST only; and in the evening, a QST and DST. Active N indicates a theoretical ceiling: the proportion of participants completing any sessions during the AP. Other participants remained enrolled but did not contribute cognitive or PRO data. A drop in participation is evident between APs 1 and 2.

**Figure 4:**
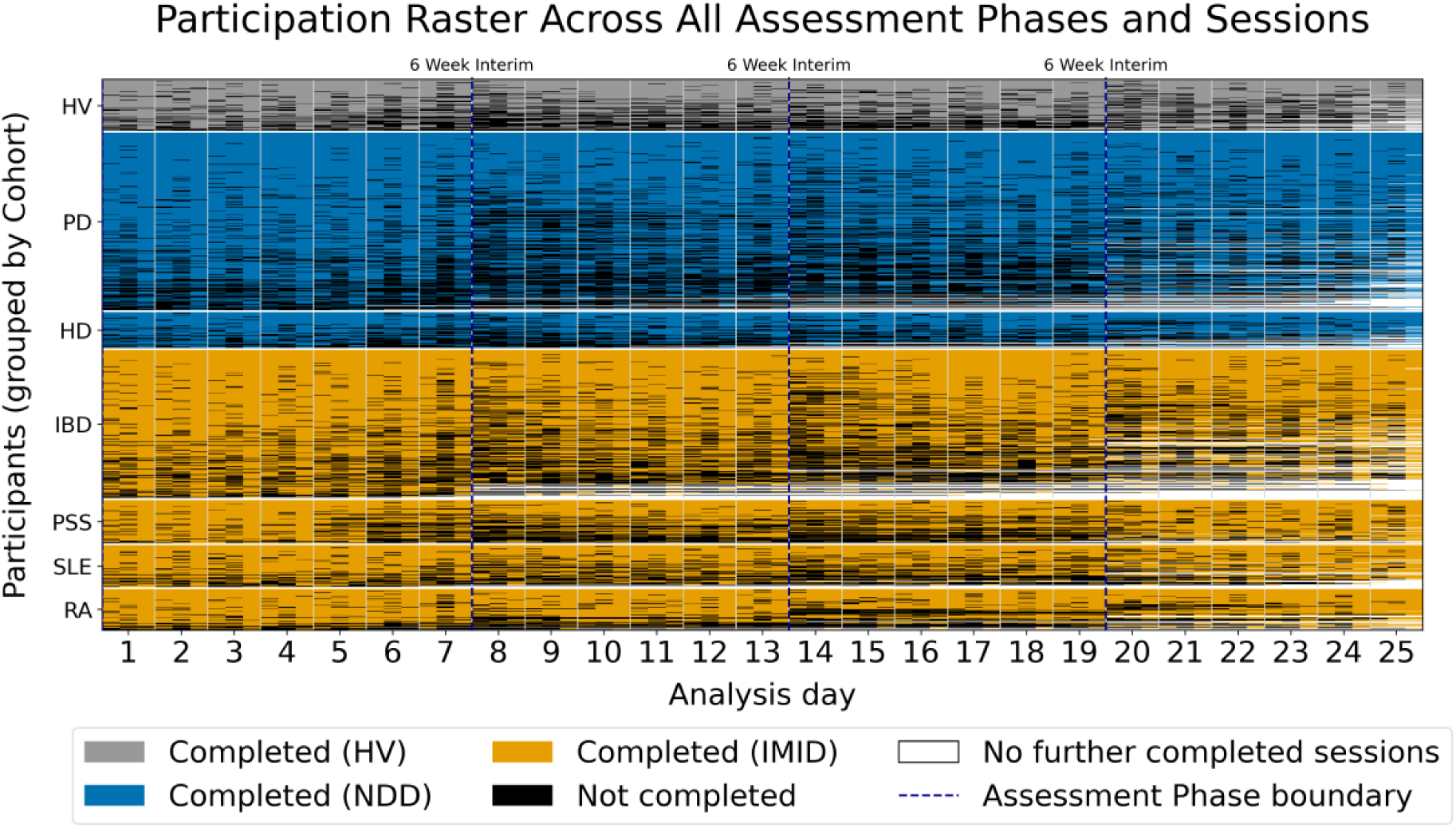
The raster shows daily participation in the three scheduled sessions, morning, afternoon, and evening, shown in order in each study day. The banding effect reflects the consistent participation pattern of evening > morning > afternoon completion, and the drop in active study participation is reflected in the increase of white shading from the start boundary of the second AP. The ‘darker’ beginnings of each AP demonstrate the pattern of increasing coverage over AP-days in APs 2, 3, and 4.

**Figure 5:**
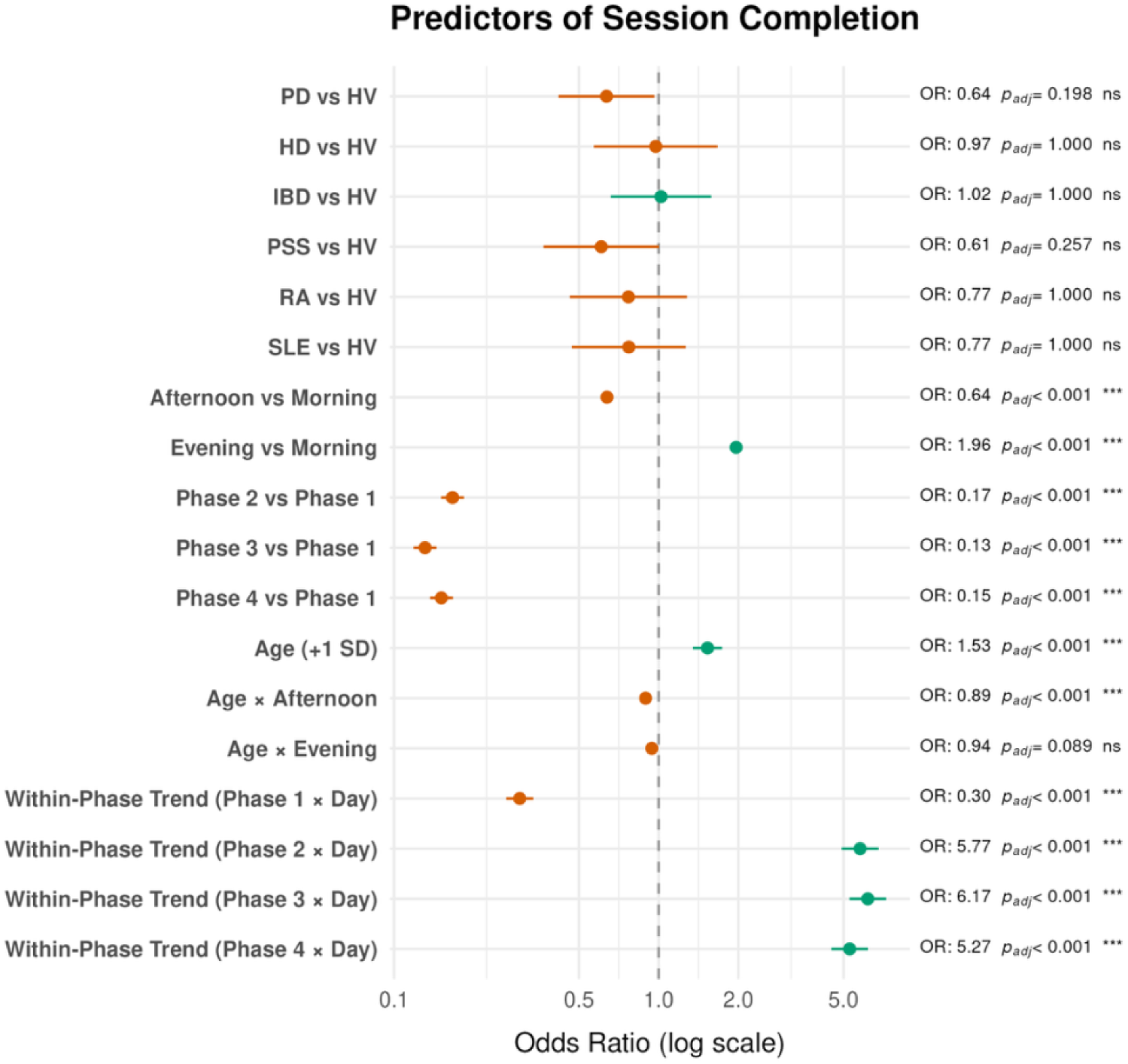
Forest plot illustrates odds ratios of fixed effects in Logistic Mixed Effects model, with 95% CI bars and corrected p-values. Where error-bars cross the 1.0 mark there is no uncorrected significant difference from reference.

**Table 3:**
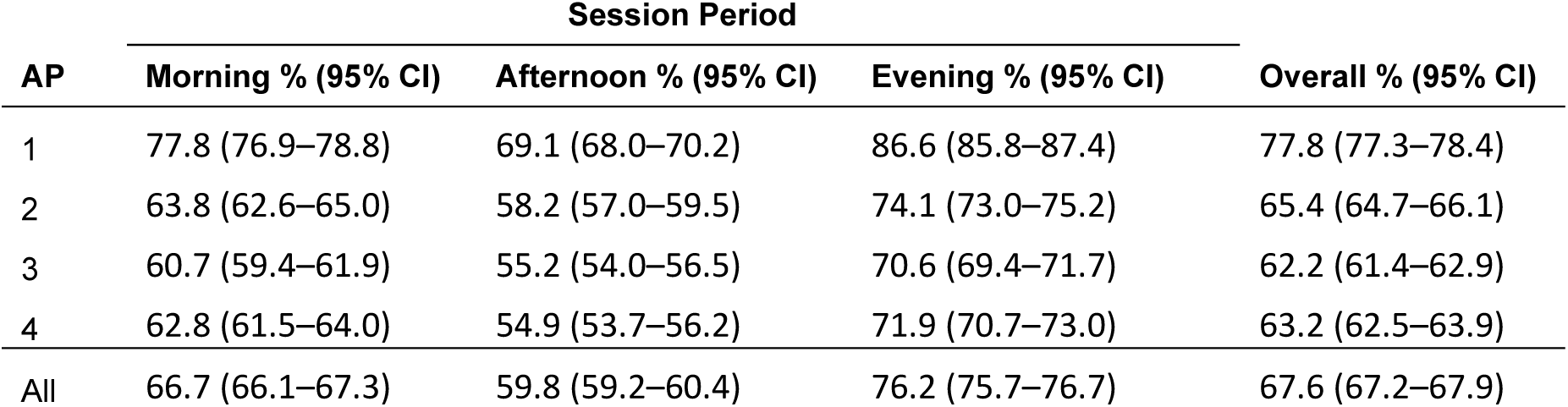
Mean observed coverage per-participant in each of the four APs, and ‘All’, across all expected sessions. The 95% CIs are ± SEM × 1.96.

The model intercept was defined as a mean-aged healthy volunteer (HV) during a morning session at the midpoint of AP 1. Full model parameter estimates are provided in Appendix Table 4.

After adjusting for age, sTOD, and site, diagnostic cohorts had no effect on coverage (all *p* > .05, Holm-corrected). The model-predicted participant adherence probabilities ranged from .653 for PSS to .760 for IBD, compared to .756 for the control HV cohort.

Coverage varied significantly by time of day (all *p* < .001, Holm-corrected). For a mean-aged participant, modelled completion probabilities were highest in the evening (.821), intermediate in the morning (.700), and lowest in the afternoon (.598). Older age was associated with higher participation across all scheduled sessions. A significant Age × sTOD interaction (*p* < .001) indicated that older participants maintained more consistent adherence across the day.

Overall coverage fell significantly following the first Assessment Phase (AP1 predicted coverage, .845), remaining lower throughout AP 2 (.685), AP 3 (.639), and AP 4 (.654), all *p* < .001 versus AP 1, Holm-corrected).

A significant interaction between AP and AP-day effects revealed a shift in within-phase trajectories. During AP 1 there was a significant decline in coverage, a predicted .911 at the start of the AP to .755 at the end. This decline reversed in subsequent APs, (all *p* < .001), with coverage instead improving from start to the end of AP 2 (.640 to .754), AP 3 (.580 to.722), and AP 4 (.621 to .721). This pattern is evident in Figure 3, above.

### 3.2. Reliability and Stability of Performance Measures

#### 3.2.1. ICC Analysis

Baseline reliability was assessed using data restricted to AP1. Results are shown in Figure 6. Reliability was generally good to excellent for the primary cognitive measures. For PVT mean reaction time, ICCs ranged from .742 (HV) to .901 (HD). DSST total correct showed similarly good consistency, ranging from a significantly lower than controls .746 in PD, to .928 (HD). DSST correct movement duration had wider variability overall, with good reliability in controls (.823) and cohorts such as RA (.822), but moderate in the PD (0.541) and PSS (0.610) cohorts. The 95% CIs for the PD and PSS cohorts did not overlap with those of the healthy control group, indicating a significant reduction in motor speed reliability for these groups.

**Figure 6:**
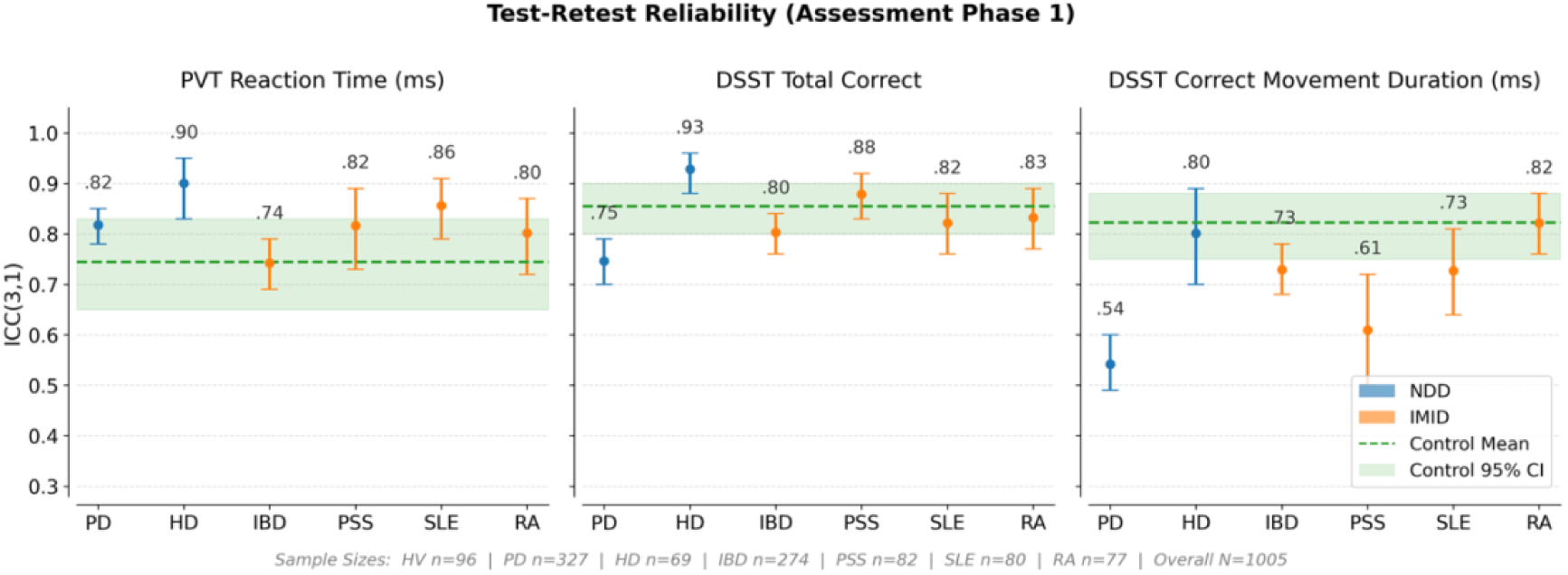
Test-retest reliability (ICC 3,1) for three cognitive performance measures. Points represent cohort means with 95% confidence intervals (CIs). Cohorts are colour-coded by disease group: neurodegenerative disease (NDD, blue), immune-mediated inflammatory disease (IMID, orange), and controls (green). The dashed green line and shaded band indicate the control mean and 95% CI. Cohorts are ordered by disease group and descending sample size.

### 3.3. Sensitivity To Age and Cognitive Impact of Disease

We used linear mixed-effects ANCOVAs, modelling age as a covariate and cohort as a fixed factor, with subject-level random intercepts to assess the tasks’ sensitivity. Age and cohort effects are illustrated in Figure 7 below.

**Figure 7:**
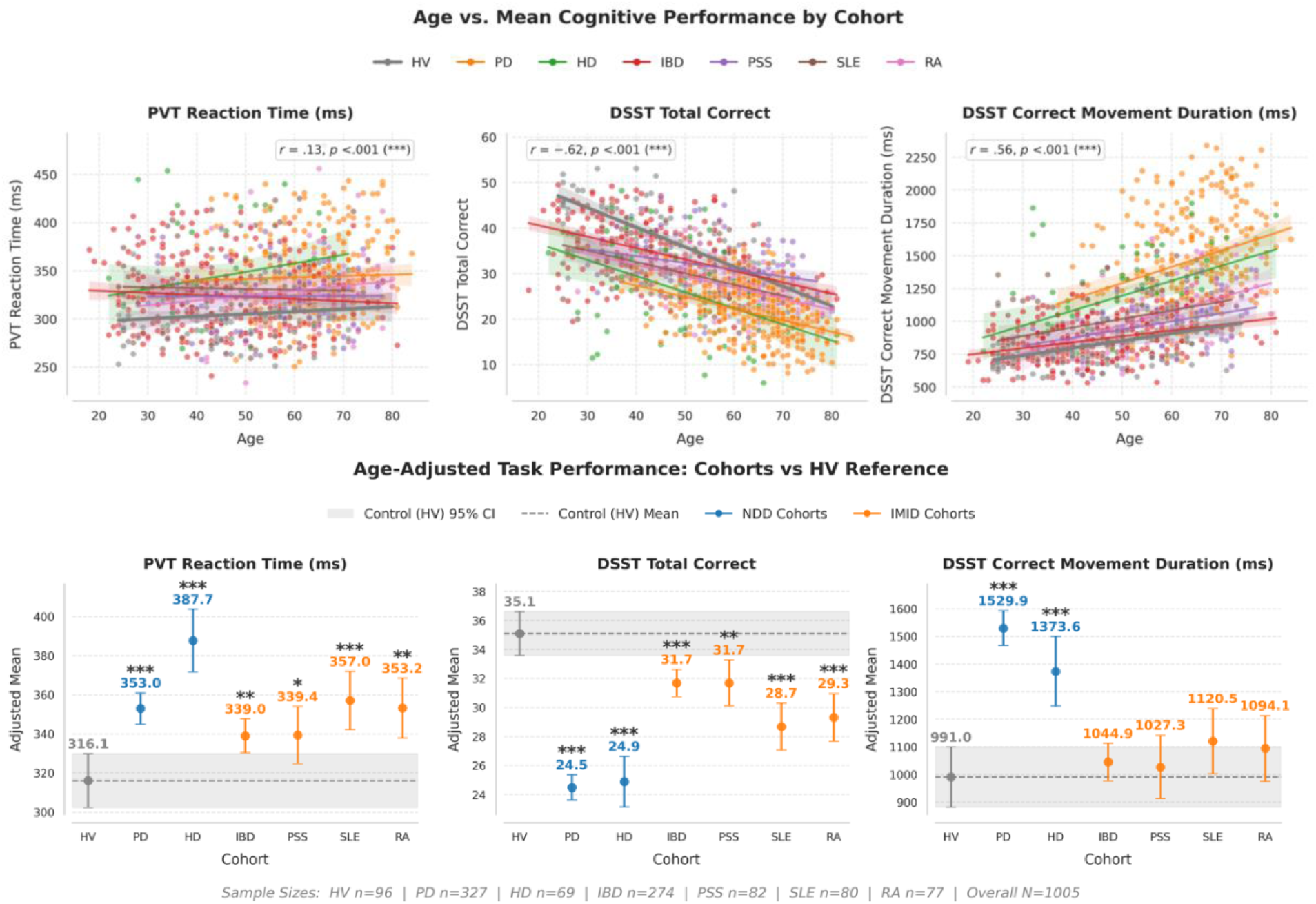
Scatter plots (top) show un-adjusted relations of PVT and DSST measures to age (with Pearson correlation annotations), and mean performance estimates from ANCOVA models for the three cognitive task outcome measures: Psychomotor Vigilance Task (PVT) mean reaction time, Digit Symbol Substitution Test (DSST) total correct responses, and DSST mean correct movement duration. Bars are estimated marginal means by cohort, adjusted for age, with 95% confidence intervals, with grey bands representing Healthy Volunteer (HV)reference group.

#### 3.3.1. PVT Mean Reaction Time

The mixed-effects ANCOVA revealed a strong overall cohort effect on mean reaction time (Wald χ²(6) = 53, *p* < .001). Relative to HVs estimated marginal mean (EMM) of 316 ms, (95% CI [305, 332]), all clinical cohorts showed significantly slower response. The HD cohort exhibited the slowest response time (*β* = +72 ms, 95% CI [51, 93], Cohen’s *d* = 0.98, *p* < .001), followed by PD (*β* = +37 ms, 95% CI [20, 54], *d* = 0.50, *p* < .001).

The IMID cohorts showed smaller but still statistically reliable lengthening of RTs compared with controls, PSS (*β* = +23 ms, 95% CI [2, 43], *p = .006), d = 0.32*, *p* = .029, RA (*β* = +37 ms, 95% CI [14, 54], *p* = .003), SLE (*β* = +41 ms, 95% CI [26, 57], *p* < .001), and IBD (*β* = +23 ms, 95% CI [8, 40], *d* = 0.31, p = .008).

Age was not significantly associated with reaction times (*β* = 0.3, 95% CI [-0.07, 0.65], *p* = .110).

#### 3.3.2. DSST Total Correct

The cohort-level model for DSST accuracy showed strong overall explanatory power (Wald χ²(6) = 208, *p* < .0001). HVs achieved an adjusted mean of 35.1 items over the 90s of the task (95% CI [33.6, 36.5]).

All cohorts performed significantly below HVs, all comparisons *p* < .001. The largest differences were in the neurodegenerative groups: PD (*β* = –10.6 items, 95% CI [–8.7, – 12.3], *d* = 0.95) and HD (*β* = –10.2 items, 95% CI [–12.5, –7.9] *d* = 0.92). All IMID cohorts showed moderate but reliable reductions: IBD (*β* = –3.4, 95% CI [–5.0, –1.6], *d = 0.31*), PSS (*β* = –3.4, 95% CI [–5.8, –2.2], *p* < .001), RA (*β* = –5.8, 95% CI [–8.3, –3.7], *d = 0.52*, *p* < .001), and SLE (*β* = –6.4, 95% CI [–8.5, –4.1], *d = 0.58*).

Age showed a negative association with DSST accuracy (*β* = –0.35 items/year, 95% CI [–0.31, –0.39], *p* < .001), reflecting the known sensitivity of DSST to age-related processing speed and executive decline.

#### 3.3.3. DSST Correct Movement Duration

The model for mean correct movement duration also showed a strong cohort effect (Wald χ²(6) = 136, *p* < .001. HVs displayed an adjusted mean movement duration of 991 ms (95% CI [883, 1104]).

As with the accuracy measure, effects in NDD cohorts were the most pronounced, with PD exhibiting the most slowing (*β* = +539 ms, 95% CI [+401, +665], *d = 0.84*) and HD also substantially slower (*β* = +383 ms, 95% CI [219, 552] , *d = 0.59*, both *p* < .001) than controls.

IMID cohorts displayed numerically longer movement durations than HV but none showed statistically significant differences following Holm correction: IBD (*β* = +54 ms, *p* = .687, *d = 0.08*), PSS (*β* = +36 ms, *d* = 0.06, *p* = .687), RA (*β* = +103 ms, *d = 0.16*, *p* = .612), SLE (*β* = +130 ms, *d* = 0.20, *p* = .458)

Age was strongly associated with slower movement durations (*β* = +12.9 ms/year, 95% CI [10.0, 15.7], *p* < .001).

### 3.4. Effects of Task Familiarisation on Performance

To characterise long-run familiarisation effects, performance change over 25 days was examined including cohort-specific interactions. These results are illustrated in Figure 8 below.

**Figure 8:**
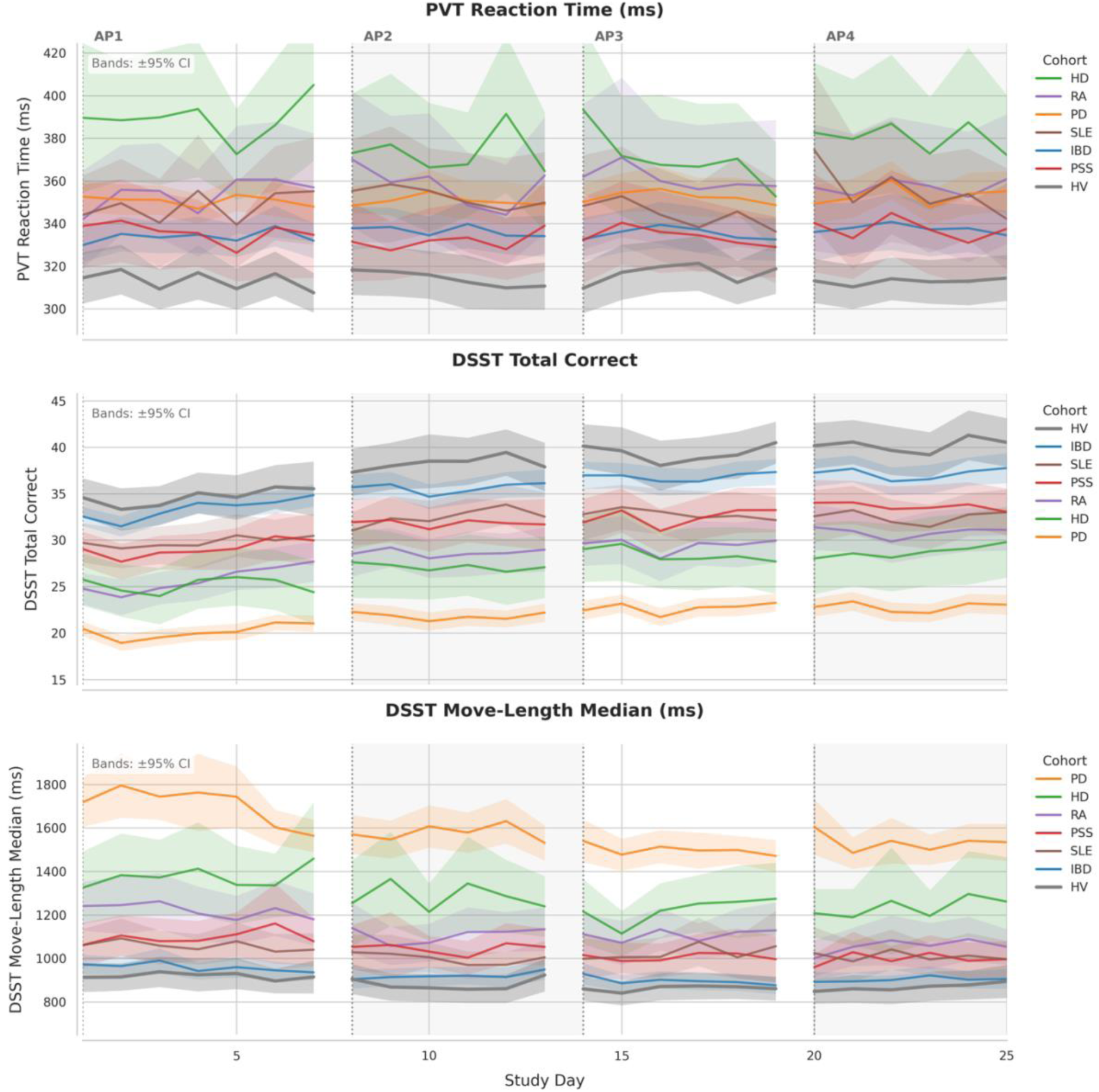
Performance in PVT and DSST over 25 study days. Vertical dashed lines indicate beginnings of APs after approximately 6-week intervals.

#### 3.4.1. PVT Mean Response Time

Across the cohorts, study-day was not a significant predictor, *p* = .44, and none of the clinical cohorts showed significant familiarisation effects (all *p* ≥ .9, Holm-corrected).

#### 3.4.2. DSST Total Correct

Familiarisation effects were evident in DSST Total Correct. The HV group improved at a rate of 0.238 items/day (*p* < .001), equating to ∼5.94 items over the 25 study days. All clinical cohorts also showed significant improvement over time, though most were significantly weaker than HV. The RA cohort improved at 0.22 items/day, ∼5.2 over the study, not significantly weaker than HV (*p* = .070). All other cohorts showed significantly reduced familiarisation compared with controls, all Holm-corrected: PSS, 0.189 items/day, 4.73 items (*p* = .017), IBD 0.177 items/day, 4.42 items (*p* < .001), SLE 0.155 items/day, 3.88 items (*p* < .001), and HD 0.159 items/day, 3.98 items (*p* < .001). The PD cohort showed the weakest familiarisation effect at 0.113 items/day, 2.83 items (*p* < .001).

#### 3.4.3. DSST Correct Movement Duration

Familiarisation effects on DSST movement duration varied across cohorts. In HV, the slope was −1.80 ms/day (*p* = .071). In contrast, two clinical cohorts showed significant familiarisation effects: the PD cohort at −7.57 ms/day (*p* < .001), and RA −6.52 ms/day (*p* = .001). The remaining cohorts showed weaker, non-significant decreases (HD −5.36 ms/day, *p* = .096, IBD: −2.87 ms/day; PSS: −2.96 ms/day; SLE: −3.01 ms/day).

## 4. Discussion

The overall assessment coverage over all active periods was 67.5% for the PVT and 77.0% for the DSST, supporting the operational feasibility of this remote testing model. These figures are similar to coverage reported in other remote paradigms utilising CANTAB tasks (e.g., the large-scale Intuition project, Butler, Yang, Brown et al., 2025) and alternative assessment batteries (e.g., Polk, Öhman, Hassenstab, et al., 2025). While initial coverage approached 90%, a subset of participants exhibited a ’silent drop-out’ effect following the first assessment period. For those retained beyond this initial drop, day-by-day session completion then remained consistent. Coverage exhibited robust, cohort-independent time-of-day effects, with completion rates lowest in the afternoon, intermediate in the morning, and highest in the evening. This Evening > Morning > Afternoon pattern mirrors findings in paradigms using repeating batteries (e.g., Thompson et al., 2024), indicating it is not driven by differing session lengths or task appeal. A significant interaction with age showed this temporal pattern was diminished in older participants, supporting the hypothesis that daytime sessions impose a higher burden on younger demographics due to more active routines.

High cohort-independent ICCs across the measures support the feasibility of using these assessments in the general population and in transdiagnostic heterogeneous populations without symptom-based bias in validity. There is a noticeable reduction in ICC for PD and PSS cohorts compared with controls and other cohorts for DSST movement duration. During the week-long baseline AP, shifts in movement symptoms in PD may alter DSST movement durations sufficiently to impact the rank-ordering of participants in PD, although this does not explain the reduced ICC in PSS. Future analyses may assess the causes of these differences.

Differences in task performance between the HV and each disease group showed that both PVT and DSST are sensitive to impacts on cognition of both neurodegenerative as well as immune-mediated inflammatory diseases. Cognitive impacts of both PD and HD are well known and substantial, and routinely captured by in-clinic rating scales such as MDS-UPDRS and MDS-UHDRS, that assess both motor and non-motor domains in PD and HD. By assessing IMID cohorts alongside cohorts with well-known benchmarks of cognitive impairment, the present study effectively contextualises the cognitive burden of IMIDs.

While PVT performance is impaired for all cohorts compared with controls, it appears markedly slow for the HD cohort (β = +72 ms), compared with PD (β = +37 ms) or the IMID cohorts (β = +22–42 ms), unlike in the DSST where PD and HD cohorts show relatively similar performance.

In our study we found significant performance effects in IMID in both PVT and DSST. Cognitive impairments have been reported and documented across several domains among IMID groups, observed with small to large effect sizes depending on the assessment and battery duration. In IBD, a meta-analysis by Hopkins et al. found modest to large deficits in attention, working memory and executive function. In PSS, Koçer *et al*. (2016) reported working memory and executive function effects, but a systematic review (Manzo *et al*., 2019) found that evidence of cognitive impairments in PSS is scarce and somewhat contradictory, and subject to issues of comorbidity, with small effect sizes. In RA, Appenzeller, Bertolo, & Costallat (2004) described a cross-sectional study that used various in-clinic paper and pencil neuropsychological measures to assess cognition and found effects on verbal fluency, memory, and working memory. Finally, in a meta-analysis of cognition in SLE, Leslie and Crowe (2018) reported significant but small effect sizes in deficits of visual attention, cognitive fluency, visual memory and visual reasoning. Across this literature, the reports of cognitive effects in these diseases have come from small studies without consistent tests. With moderate effect sizes, the results of our study show that two briefly administered tasks are sensitive to diverse impacts on cognition of IMID.

Beyond static performance, the longitudinal trajectories of these metrics may provide additional clinical insights. In analysis of longitudinal effects, early familiarisation effects could have been missed in the first three onboarding days of AP 1 that were not analysed. However, over the course of the study, as expected for a simple task assessing RT, the PVT did not exhibit familiarisation-based improvements, and RTs remained stable over the study duration. Conversely, significant familiarisation effects were observed in the DSST. This aligns with prior literature (e.g., Jaeger et al., 2018) that attributes such improvements to familiarisation with the testing interface and stimuli, rather than the memorisation of digit-symbol mappings, which vary between visits. Among healthy volunteers, DSST performance improved by approximately 6 correct answers over the four assessment phases from a baseline average in the mid-thirties. This familiarisation effect was blunted in all disease cohorts except RA, and was weakest in the PD cohort, where the improvement was roughly half that of the healthy volunteers. This differential capacity to learn the task may warrant further investigation, as attenuated familiarisation effects may themselves serve as a sensitive marker of cognitive impairment. The differentiation and contextualisation of these measures over diverse cohorts may in future work help establish their sensitivity in distinguishing known-group impairments. Subsequent analyses of these study data will examine the relations of these objective cognitive measures to participants’ subjectively-reported changes of state, as well as relations of the measures to clinical assessments of disease severity.

## 5. Conclusions

This study showed that in-home high frequency testing is feasible at scale and broadly acceptable across a clinically diverse population with varying cognitive and motor disease burdens. Extending prior work in separate disease cohorts, this suggests smartphone-based assessments can be reliably and repeatedly deployed across multiple chronic conditions in patients’ own environments.

The cognitive tasks were sensitive to the known impacts of disease, providing objective digital markers that distinguished control and patient cohorts. Differences in familiarisation trajectories over the study suggest cohort-specific patterns not captured in conventional clinic-based testing. However, the primary utility of study methods such as those explored here lie in the ability to monitor changes in cognitive health or disease progression during the ordinary course of daily living. These findings support the viability of such testing methods for a variety of purposes.

## Data Availability

All data produced in the present study are available upon reasonable request to the authors

## Acknowledgement

The IDEA-FAST project has received funding from the Innovative Medicines Initiative 2 Joint Undertaking under grant agreement No. 853981. This Joint Undertaking receives support from the European Union’s Horizon 2020 research and innovation programme and EFPIA and associated partners.

This communication reflects the view of the authors and neither IMI nor the European Union and EFPIA are liable for any use that may be made of the information contained herein.

## Footnotes

a The study has received funding from the Innovative Medicines Initiative 2 Joint Undertaking under grant agreement No 853981.

## 8 Appendix

**Table 4:** Parameter estimates from the logistic mixed model of assessment coverage. P-value adjustment uses Holm’s method.

| Family | Term | Logit [95% CI] | SE | z | p | p (adj.) | OR [95% CI] |
| --- | --- | --- | --- | --- | --- | --- | --- |
| **Baseline** | Baseline log-odds (AP 1, sTOD Morn, HV, mean age) | 2.438 [2.012, 2.864] | 0.217 | 11.22 | <0.001 | — | 11.45 [7.48, 17.53] |
| **Session Time-of-Day** | A vs Morning | -0.450 [-0.494, -0.406] | 0.023 | -19.94 | <0.001 | <0.001 | 0.64 [0.61, 0.67] |
| Session Time-of-Day | E vs Morning | 0.674 [0.626, 0.721] | 0.024 | 27.91 | <0.001 | <0.001 | 1.96 [1.87, 2.06] |
| **Age** | Age (per SD) at sTOD Morning | 0.425 [0.298, 0.553] | 0.065 | 6.54 | <0.001 | <0.001 | 1.53 [1.35, 1.74] |
| **Age × sTOD** | Age × sTOD A (diff vs Morn) | -0.114 [-0.158, -0.069] | 0.023 | -5.04 | <0.001 | <0.001 | 0.89 [0.85, 0.93] |
| Age × sTOD | Age × sTOD E (diff vs Morn) | -0.060 [-0.107, -0.013] | 0.024 | -2.49 | 0.013 | 0.089 | 0.94 [0.90, 0.99] |
| **AP number** | AP 2 vs AP 1 | -1.794 [-1.894, -1.694] | 0.051 | -35.21 | <0.001 | <0.001 | 0.17 [0.15, 0.18] |
| AP number | AP 3 vs AP 1 | -2.033 [-2.133, -1.934] | 0.051 | -40.02 | <0.001 | <0.001 | 0.13 [0.12, 0.14] |
| AP number | AP 4 vs AP 1 | -1.891 [-1.990, -1.791] | 0.051 | -37.18 | <0.001 | <0.001 | 0.15 [0.14, 0.17] |
| **AP day** | AP-day slope in AP 1 | -1.209 [-1.327, -1.091] | 0.06 | -20.12 | <0.001 | <0.001 | 0.30 [0.27, 0.34] |
| **AP × AP-day** | AP 2 × day (diff vs AP 1) | 1.753 [1.593, 1.913] | 0.082 | 21.43 | <0.001 | <0.001 | 5.77 [4.92, 6.78] |
| AP × AP-day | AP 3 × day (diff vs AP 1) | 1.820 [1.661, 1.979] | 0.081 | 22.4 | <0.001 | <0.001 | 6.17 [5.26, 7.24] |
| AP × AP-day | AP 4 × day (diff vs AP 1) | 1.662 [1.503, 1.821] | 0.081 | 20.44 | <0.001 | <0.001 | 5.27 [4.49, 6.18] |
| **Cohort** | HD vs HV | -0.026 [-0.563, 0.512] | 0.274 | -0.09 | 0.926 | 1.000 | 0.97 [0.57, 1.67] |
| Cohort | IBD vs HV | 0.020 [-0.418, 0.458] | 0.223 | 0.09 | 0.927 | 1.000 | 1.02 [0.66, 1.58] |
| Cohort | PD vs HV | -0.454 [-0.871, -0.037] | 0.213 | -2.13 | 0.033 | 0.198 | 0.64 [0.42, 0.96] |
| Cohort | PSS vs HV | -0.499 [-1.002, 0.003] | 0.256 | -1.95 | 0.051 | 0.257 | 0.61 [0.37, 1.00] |
| Cohort | RA vs HV | -0.263 [-0.774, 0.248] | 0.261 | -1.01 | 0.313 | 1.000 | 0.77 [0.46, 1.28] |
| Cohort | SLE vs HV | -0.260 [-0.755, 0.236] | 0.253 | -1.03 | 0.304 | 1.000 | 0.77 [0.47, 1.27] |

